# Post-traumatic Stress Risk among COVID-19 Survivors in Colombia

**DOI:** 10.1101/2021.12.02.21267210

**Authors:** Adalberto Campo-Arias, John Carlos Pedrozo-Pupo, Edwin Herazo

## Abstract

The study’s objective was to establish the prevalence and variables associated with post-traumatic stress disorder risk (PTSD-R) in a sample of COVID-19 survivors in Santa Marta, Colombia. A cross-sectional study was designed with a non-probabilistic sample of adult COVID-19 survivors. Participants were demographically characterized and completed scales for depression risk, insomnia risk, and PTSD-R. Three hundred and thirty COVID-19 survivors between 18 and 89 years participated; 61.52% were women. The frequency of depression risk was 49.70%; insomnia risk, 60.61%; and PTSD-R, 13.33%. Depression risk (OR = 41.43, 95% CI 5.54 – 311.63), insomnia risk (OR = 5.25, 95% CI 1.77 – 18.71), low income (OR = 3.46, 95% CI 1.38 – 8.67) and being married or free union (OR = 2.65, 95% CI 1.13 – 6.22) were associated with PTSD-R. In conclusion, two out of every fifteen COVID-19 survivors are in PTSD-R. Depression and insomnia risk are strongly associated with PTSD-R in COVID-19 survivors.

## Introduction

Post-traumatic stress symptoms are frequent in people who have lived a life experience that significantly threatened their physical or psychological integrity. Traumatic events include military combat, violent assaults, physical or sexual abuse, natural disasters, and serious accidents. Post-traumatic stress disorder (PTSD) is characterized by intrusive symptoms (distressing memories or dreams), avoidance of situations related to the stressful event (people, places, or conversations), cognitive and mood disturbances (persistent negative beliefs or negative emotions), and reactivity (irritability, hypervigilance or sleep disturbances) (American Psychiatric Association, APA, 2013).

During the COVID-19 pandemic, high numbers of people from the general population (Karaivazoglou et al., 2021), healthcare workers (Chatzittofis et al., 2021), and survivors of infection are at high risk of post-traumatic stress disorder (PTSD-R) (Bellan et al., 2021; Cai et al., 2020; Chang & Park, 2020; De Lorenzo et al., 2020; Einvik et al., 2021; Liu et al., 2020; Matalon et al., 2021; Poyraz et al., 2021; Simani et al., 2021; Tarsitani et al., 2021).

The prevalence of PTSD-R is between 5.8% and 34.5% in COVID-19 survivors, depending on the population’s characteristics and measurement instrument. The PTSD-R, measured with the Post-Traumatic Stress Disorder Checklist-5 (PCL-5), in China, Chang et al. (2020) observed in 64 post-hospitalization survivors that 20.3%. In Norway, Einvik et al. (2021), out of 1,149 participants, found that 7.4% scored for PTSD-R. In Italy, Tarsitani et al. (2021), in 115 survivors three months after hospital discharge, 10.4% were found to have PTSD-R. In China, Liu et al. (2020), in a sample of 675 survivors, including 90 doctors and nurses, found that 12.2% scored for PTSD-R. In Iran, Simani et al. (2021) documented a prevalence of 5.8% six months post-hospitalization. Likewise, the PTSD-R, explored with the Impact of Event Scale-Revised, in Italy, Bellan et al. (2021), in 238 survivors of severe COVID-19, observed that 17.2% were in PTSD-R four months after hospital discharge. In Italy, De Lorenzo et al. (2021), in 185 participants found that 22.2% had TERP three to four weeks after leaving the hospital. In Turkey, Poyraz et al. (2021), in 98 survivors approximately two months after discharge, 34.5% reported PTSD-R.

Variables associated with PTSD-R in COVID-19 survivors are divergent findings. The prevalence of PTSD-R has been observed to be independent of age in research (Chang & Park, 2020; Tarsitani et al., 2021); however, others found that the frequency increased with age (Cai et al., 2020; Liu et al., 2020). Likewise, some studies show that the frequency of PTSD-R is higher in women (Bellan et al., 2021; Einvik et al., 2021; Tarsitani et al., 2021); however, a more significant number reported no differences by gender (Cai et al., 2020; Chang & Park, 2020; Liu et al., 2020; Poyraz et al., 2021; Simani et al., 2021).

In the same sense, a more significant number of cases have been documented in more symptomatic survivors or with greater severity of the infection (Einvik et al., 2021; Liu et al., 2021) and more days of hospital stay (Matalon et al., 2021; Tarsitani et al., 2021). However, a lack of relationship between the length of hospitalization and PTSD-R has been reported (Chang & Park, 2020), with even shorter hospital stay with tests being related to PTSD-R (Liu et al., 2020).

In addition, it has been shown that the PTSD-R is higher in the presence of comorbidity (Tarsitani et al., 2021), symptoms for more extended periods (Poyraz et al., 2021), the persistence of residual symptoms (Cai et al., 2020), and symptoms of depression (Matalon et al., 2021; Poyraz et al., 2021) or insomnia (Poyraz et al., 2021). The relation between treatment in the intensive care unit and PTSD-R is inconsistent (Liu et al., 2020; Tarsitani et al., 2021), and no differences were observed in the prevalence of PTSD-R between health workers and the general population (Liu et al., 2020), outpatient or in-hospital treatment (Einvik et al., 2021), and the discharge time (Chang & Park, 2020).

In Colombia, by December 2nd, 2021, the COVID-19 pandemic has left 5,074,079 confirmed cases, 35,515 active cases, 4,913,921 survivors, and 128,643 deaths [rate of 2,491 deaths per million inhabitants] (Ministry of Health and Social Protection of Colombia, 2021). Consequently, a high number of COVID-19 survivors can be expected to be in PTSD-R. General practitioners who serve a large part of the demand for mental health care in low- and middle-sized countries income such as Colombia should actively investigate symptoms of the PTSD spectrum (Campo-Arias & Barliza, 2020; Montes-Arcón & Campo-Arias, 2020). COVID-19 survivors can complete scales to screen PTSD cases in the waiting room (Colston et al., 2016). Healthcare professionals must confirm the diagnosis of PTSD using a clinical interview (APA, 2013) and initiate the management of most of what s cases (Campo-Arias & Barliza, 2020). PTSD negatively affects daily activities (APA, 2013), enjoyment of life (Jind, 2001) and increases the risk of suicidal behaviors (Kim & Thomas, 2019).

This study analyzes a sample of Colombian Caribbean COVID-19 survivors. A region of the country is characterized by a high frequency of familism, psychological and financial support from relatives, neighbors, and friends (Krysinska & Lester, 2010). In addition, the relationship between depression risk, insomnia risk, and PTSD-R are explored. Previous studies that simultaneously measured these outcomes have failed to adjust for the possible confounding effect that these variables can generate (Cai et al., 2020; Liu et al., 2020). Depression and insomnia are part of symptoms for PTSD diagnosis (APA, 2013).

The study’s objective was to establish the prevalence and variables associated with PTSD-R in a sample of COVID-19 survivors in Santa Marta, Colombia.

## Methods

### Design and sample

A cross-sectional analytical study was designed. There was a convenience sample of COVID-19 survivors who physically or virtually consulted a pulmonology service for symptoms related or not to COVID-19. To establish a prevalence of 25% (± 5), with a confidence level of 95%, based on other studies that showed prevalence between 5% and 35% (Bellan et al., 2021; Cai et al., 2020; Chang & Park, 2020; De Lorenzo et al., 2020; Einvik et al., 2021; Liu et al., 2020; Matalon et al., 2021; Poyraz et al., 2021; Simani et al., 2021; Tarsitani et al., 2021). A sample of 288 participants had to be completed (Hernández, 2006). Likewise, associations with acceptable 95% confidence intervals (95% CI) could be estimated (Katz, 2006).

### Instruments

#### Depression risk

Depression risk was assessed with the Patient Health Questionnaire (PHQ-9). The PHQ-9 quantifies symptoms of depression during the previous two weeks with nine items of four response options, which are scored from zero to three (Kroenke et al., 2001). In the present sample, the PHQ-9 presented Cronbach’s alpha of .85. For Colombia, 7 ≤ indicate depression risk, with a sensitivity of .90, specificity of .82, and Cronbach’s alpha of .80 (Cassiani-Miranda et al., 2021).

#### Insomnia risk

Insomnia risk was quantified with the Athens Insomnia Scale (AIS). The AIS brings together eight items with response options that are scored from zero to four. 6 ≤ risk of insomnia is considered (Soldatos et al., 2000). In the present sample, the AIS reached Cronbach’s alpha of .85. In Colombia, the AIS has shown a Cronbach’s alpha of .93 (Campo-Arias et al., 2020).

#### PTSD-R

The PTSD-R was measured with the SPAN. The SPAN consists of four items with five response options scored from one to five (Meltzer-Brody et al., 1999). Scores ≥ 12 were classified as PTSD-R based on a Colombian study that used the same cut-off point; In this research, the SPAN showed Cronbach’s alpha of .81 (Pedrozo-Pupo & Campo-Arias, 2020). The SPAN presented Cronbach’s alpha of .81 in the present sample.

### Procedure

The patients were contacted in the pulmonology outpatient service of three institutions. Patients were informed of the study objectives of giving informed consent. Patients completed the research questionnaire an online link between October 12, 2020, and April 30, 2021.

### Statistical analysis

In the descriptive analysis, frequencies and percentages were observed for categorical variables, and mean (M), and standard deviation (SD) were calculated for quantitative variables. PTSD-R was considered the dependent variable, and the remaining variables were treated as independent variables. Crude and adjusted odds ratios (OR) and 95% CI were estimated. To avoid overestimation for ORs with a wide 95% CI, additional log ORs were calculated. This alternative is recommended when the prevalence of the dependent variable is greater than 10% (Bastos et al., 2015).

Greenland’s recommendations were considered to adjust for significant associations. The first recommendation is to test the effect of all variables that show probability values ≥ .20. The second is to retain the variables that show significant associations during the adjustment process. Furthermore, the third is to leave in the final model the non-significant variables that induce a variation ≥ 10% in the association that presents the highest OR (Hosmer et al., 1991). Additionally, the final model had to fit adequately, showing a Hosmer-Lemeshow test with a p-value > .10 (Greenland, 1989). The analysis was completed in the Project Janovi 1.8.2 program.

### Ethical considerations

A research ethics committee approved the study of a Colombian university (Approval Certificate 002 of March 26, 2020). Participants signed informed consent voluntarily and without receiving incentives. The instruments used in the research are free to use. The study followed the Colombian standard in health research (Ministerio de Salud de Colombia, 1993) and the international regulation for the participation of humans in research (World Medical Association, 2018).

## Results

Three hundred and thirty thee COVI-19 survivors were invited to participate; 2.08% (n = 7) rejected the invitation. The 330 participants were between 18 and 89 years old (Mean = 47.67, SD = 15.17), 61.52% were women, 62.42% had university studies, 66.06% were married or union-free 71.21% with low family income. The prevalence of risk of depression was 49.70%; risk of insomnia, 60.61%; and PTSD-R, 13.33%. More characteristics of the population are presented in Table 1.

**Table 1.** Sample description (N = 330).

| Variable | Category | n | % |
| --- | --- | --- | --- |
| Age (years) | 18 – 59 | 263 | 79.70 |
|  | 60 or more | 67 | 20.30 |
| Gender | Female | 203 | 61.52 |
|  | Male | 127 | 38.48 |
| Education | Primary | 29 | 8.79 |
|  | Secondary | 95 | 28.79 |
|  | University | 206 | 62.42 |
| Family income | Low | 235 | 71.21 |
|  | High | 95 | 28.79 |
| Married or civil partnership | Yes | 218 | 66.06 |
|  | No | 112 | 33.94 |
| Healthcare worker | Yes | 47 | 14.24 |
|  | No | 283 | 85.76 |
| Comorbidity | Yes | 122 | 37.97 |
|  | No | 208 | 63.03 |
| Severity | No symptoms or mild | 218 | 66.06 |
|  | Moderate or severe | 112 | 33.94 |
| No symptoms or less three weeks | Yes | 264 | 80.00 |
|  | No | 66 | 20.00 |
| Remission lower than two months | Yes | 214 | 70.03 |
|  | No | 89 | 29.97 |
| Depression risk | Yes | 164 | 49.70 |
|  | No | 166 | 50.30 |
| Insomnia risk | Yes | 200 | 60.61 |
|  | No | 130 | 39.39 |
| PTSD – R* | Yes | 44 | 13.33 |
|  | No | 286 | 86.67 |

\* Post-traumatic stress disorder risk.

The crude ORs showed a statistically significant association between depression risk, insomnia risk, age under 60 years, low family income, married or living in a free union, moderate or severe infection, symptoms ≤ three weeks, and PTSD-R. See the OR and 95% CI values in Table 2.

**Table 2.** Crude association for PTSD-R in COVID-19 survivors.

| Variable | OR (95% CI) |
| --- | --- |
| Age between 18 and 59 | 3.94 (1.18 – 13.14) |
| Female gender | 1.58 (0.79 – 3.15) |
| Primary or secondary education | 1.18 (0.62 – 2.25) |
| Low income | 2.35 (1.01 – 5.47) |
| Married or civil partnership | 2.19 (1.01 – 4.73) |
| Healthcare worker | 0.75 (0.28 – 1.99) |
| Comorbidity | 1.50 (0.79 – 2.85) |
| Moderate or severe COVID-19 | 2.42 (1.28 – 4.61) |
| Symptoms more than three weeks | 2.50 (1.25 – 5.01) |
| Remission inferior to two months | 0.68 (0.34 – 1.33) |
| Depression risk | 58.64 (7.93 – 431.53)<sup>1</sup> |
| Insomnia risk | 10.92 (3.30 – 36.07)<sup>2</sup> |

<sup>1</sup>Log OR = 4.11 (95% CI 2.15 – 6.13).
<sup>2</sup>Log OR = 2.43 (95% CI 1.23 – 3.64).

In addition, the gender variable was considered for adjustment, which showed an association with a p-value of .19. After adjusting, depression risk, insomnia risk, a low family income, and being married or living in a free union kept the association estimates within the significant range, with adequate goodness of fit (Hosmer-Lemeshow test’s chi-squared of 1.35, degrees of freedom of 7 and p-value of .99). These variables are presented in Table 3.

**Table 3.** Adjusted association for PTSD-R in COVID-19 survivors.

| Variable | Adjusted OR (95% CI) |
| --- | --- |
| Depression risk | 41.53 (5.54 – 311.63)<sup>1</sup> |
| Insomnia risk | 5.25 (1.47 – 18.71)<sup>2</sup> |
| Low income | 3.46 (1.38 – 8.67) |
| Married or civil partnership | 2.65 (1.13 – 6.22) |

<sup>1</sup>Log odds ratio = 3.72 (95% CI 1.71 – 5.73).
<sup>2</sup>Log odds ratio = 1.74 (95% CI 1.52 – 2.94).

## Discussion

Even in a cultural context with high familism and social support, 13% of COVID-19 survivors present PTSD-R; the risk is significantly higher in the presence of risk of depression, risk of insomnia, low family income, and having a stable partner in COVID-19 survivors.

The prevalence of PTSD-R varies widely in COVID-19 survivors. In the present study, it was observed that 13% of the participants are in PTSD-R. This observation is consistent with research conducted in China (Liu et al., 2020) and Italy (Tarsitani et al., 2021), which showed comparable values between 10% and 12%. However, lower frequencies were observed in survivors in Iran (Simani et al., 2021) and Norway (Einvik et al., 2021), with prevalences between 6% and 8%, and higher in China (Cai et al., 2020; Chang & Park, 2020), Israel (Matalon et al. 2021), Italy (Bellan et al., 2021; De Lorenzo et al., 2020) and Turkey (Poyraz et al., 2021), frequencies between 17% and 35%. These disparities in the observed frequencies, even in studies carried out in the same country, can be explained by the characteristics of the population, the sampling design, and the measurement instruments (Marinova & Maercker, 2015).

In the present sample of COVID-19 survivors, age and PTSD-R were not observed to be related. This data is consistent with other investigations that showed the frequency of PTWR was similar in different age groups (Chang & Park, 2020; Tarsitani et al., 2021). However, other studies have reported that PTWT is more prevalent in older ages (Cai et al., 2020; Liu et al., 2020).

The relationship between gender and PTSD-R in COVID-19 survivors is inconsistent. The finding of the present study is consistent with several studies that reported that the prevalence of TTPN is similar in men and women (Cai et al., 2020; Chang & Park, 2020; Liu et al., 2020; Poyraz et al., 2021; Simani et al., 2021). However, other research has documented that the prevalence of PTWT is usually higher in women than in men, similar to what was observed before the COVID pandemic (Bellan et al., 2021; Einvik et al., 2021; Tarsitani et al., 2021).

Among the predisposing factors for PTSD, there are biological and socioeconomic elements. In COVID-19 survivors, the relationship between marital status, family income, and PTSD-R had not been reported. It is possible to think that people in a stable relationship, generally with children and low family income, have a higher PTSD-R. Greater vulnerability to PTSD has been observed in people who simultaneously face various psychosocial stressors (Bryant, 2019; Marinova & Maercker, 2015).

It is plausible that the characteristics of the COVID-19 infection and treatment’s circumstances are related to PTSD-R. The current study shows that infection characteristics, healthcare workers, and comorbidity were independent of PTSD-R after adjusting for other variables. This observation is not novel. It has been documented that TTPN is independent of the place of treatment (outpatient or in-hospital) (Einvik et al., 2021), the duration of hospitalization for COVID-19 (Chang & Park, 2020), or the management in the care unit intensive (Liu et al., 2020; Tarsitani et al., 2021) and discharge time (Chang & Park, 2020). Likewise, contrary to expectations, it has been reported that the PTSD may be higher when the hospital stay is shorter (Liu et al., 2020), and health workers present the same PTSD as the general population (Liu et al., 2020). However, other researchers have observed that PTSD-R is greater depending on the number and severity of symptoms (Einvik et al., 2021; Liu et al., 2020; Poyraz et al., 2021), the longer length of hospital stay (Matalon et al., 2021; Tarsitani et al., 2021), comorbidity (Tarsitani et al., 2021), and persistence of residual symptoms (Cai et al., 2020).

To date, the association between depression, insomnia, and PTSD-R has been little explored in COVID-19 survivors. In the present investigation, the risk of depression and insomnia showed a strong association with PTSD-R. This finding is consistent with previous studies that showed that symptoms of depression (Matalon et al., 2021; Poyraz et al., 2021) and manifestations of insomnia were persistent in COVID-19 survivors (Poyraz et al., 2021). This observation is consistent with the diagnostic criteria for PTSD that include changes in mood and sleep pattern (APA, 2013).

The variables associated with PTSD-R may vary due to the design of the studies and the social determinants of mental health (Álvarez-Hernández & Delgado-De la Mora, 2015). The risk factors for PTSD converge predisposing or proximal vulnerabilities (genetic and physical factors, lifestyle and social and community networks), intermediate (living and working conditions), and distal (socioeconomic, cultural, and environmental conditions) (Bryant, 2019; Marinova & Maercker, 2015).

### Practical implications

Symptoms of PTSD are common in critically ill survivors, between 15% and 45% of patients in intensive care units six to twelve months after discharge (Parker et al., 2015). By July 8, 2021, around the world, the COVID-19 pandemic has affected more than 186 million people and 4 million deaths (Ministry of Health and Social Protection of Colombia, 2021). This case-fatality rate explains PTSD symptoms in COVID-19 survivors due to fear or imminence of death. Similarly, confinement can be configured as a stressor and not necessarily a formal traumatic event (Soloveva et al., 2021).

Primary care professionals play a vital role in identifying and managing psychological symptoms in COVID-19 survivors (Campo-Arias & Barliza, 2020; Montes-Arcón & Campo-Arias, 2020). For the diagnosis of PTSD, the presence of fear of infection is insufficient; it is always necessary to specify the exposure to one or more events that involved real death threats related to the pandemic (APA, 2013). It is crucial to reduce the PTSD-R in people diagnosed with COVID-19 through preventive actions such as first aid and psychological and cognitive-behavioral support for and family members, according to particular needs (Soloveva et al., 2021).

### Study strengths and limitations

This research has the strength of showing the frequency and variables associated with PTSD-R in an adequate sample of COVID-19 survivors in the socio-cultural context of the Colombian Caribbean (Hernández, 2006; Katz, 2006). However, it has the limitation that the findings cannot be generalized due to non-probabilistic sampling (Álvarez-Hernández & Delgado-De la Mora, 2015). In the same way, the psychological variables were quantified with measurement scales that only screen and can overestimate the prevalence because they only indicate people at high risk of meeting criteria for a formal disorder (Álvarez-Hernández & Delgado-De la Mora, 2015). The diagnosis of a mental disorder is a complex clinical process that considers the presence of symptoms, the deterioration in global functioning or quality of life, and the factors that can explain the clinical manifestations, for example, comorbidity, drug use, substance abuse that induces dependence and normative events such as grief (APA, 2013).

### Conclusions

It is concluded that two out of every fifteen COVID-19 survivors are in PTSD-R. Depression and insomnia risks are strongly associated with PTSD-R in survivors in Santa Marta, Colombia. Early identification of PTSD-R and management is essential. Research is needed to evaluate the long-term PTSD-R in a probability sample.

## Data Availability

The data that support the findings of this study are available from the corresponding author upon reasonable request.

## Acknowledgment of conflicts of interest

The authors have no conflicts of interest to declare.

